# A randomized, phase IIa treatment delayed-start trial of the oral JAK 1/2 inhibitor, baricitinib, in adult idiopathic inflammatory myopathy

**DOI:** 10.64898/2026.07.30.26359332

**Authors:** Ashma Krishan, Luke Tomlinson, James B. Lilleker, Gema Sylvestre Garcia, Andrew Snedden, Mohammed Zubair, Patrick Gordon, Athiveeraramapandian Prabu, Sarah Tansley, Aamir Aslam, Helene Alexanderson, Ingrid E. Lundberg, Janine A. Lamb, Hector Chinoy

## Abstract

**Objectives:** To assess the effects of 24 weeks active treatment with baricitinib, a JAK1/2 inhibitor, in adult idiopathic inflammatory myopathy (IIM).

**Methods:** Patients with active dermatomyositis (DM) or polymyositis (PM) were enrolled into a 1:1 randomized treatment delayed-start design clinical trial (NCT04208464). Participants received 24 weeks baricitinib plus 12 weeks follow-up (Immediate-start), or 12 weeks standard of care plus 24 weeks baricitinib (Delayed-start). The primary outcome was clinical response after 24 weeks active treatment, defined as minimal improvement (Total Improvement Score >20 [TIS20]). Secondary outcomes included: TIS40 (moderate), TIS60 (major) response, between-arm comparison, time to achieve response, change in clinical outcome measures. steroid-sparing and cumulative adverse events.

**Results:** 14/15 (93%) randomized participants (mean age 43.2 years [11.6 SD]; 13 DM, 2 PM; 11 female) completed the study (baseline to 36 weeks) and all achieved TIS20 at 24 weeks post-active treatment (95% exact CI 0.68-1.00). 9/15 (60%) achieved TIS40 response and 2/15 (13%) TIS60 response. At 12 weeks post-randomization, 11/15 (73%) patients achieved at least TIS20 (95%CI 0.45-0.92), including all Immediate-start arm patients and four Delayed-start arm patients. At the same time point, evidence of a difference was noted for patient global, extramuscular, CDASI skin activity, pain, fatigue and SF-36 mental/physical health scores. Two hospitalisation serious adverse events were documented, neither related to study drug.

**Conclusions:** Treatment of IIM with baricitinib resulted in improved clinical outcome after 24 weeks. Significant improvement after 12 weeks treatment was also evident. No significant safety concerns were raised. A randomized placebo-controlled trial is needed to confirm the efficacy in patients with IIM.

**Clinical trial registration:** EudraCT Number: 2019-003868-42

ISRCTN Number /Clinical trials.gov Number: NCT04208464

https://clinicaltrials.gov/study/NCT04208464

## Introduction

Myositis, or the idiopathic inflammatory myopathies (IIM) are a group of rare autoimmune diseases of considerable health significance affecting both adults and children, and negatively impacting the lives of an estimated 10,000 participants in the UK and 250,000 worldwide^1^. The hallmark of IIM is inflammation of muscle tissue, myositis, which can lead to profound weakness, fatigue and disability. IIM is a heterogenous disease that based on clinical, serological and histopathological differences can be defined into the major clinical subgroups: dermatomyositis (DM), polymyositis (PM), inclusion body myositis (IBM), antisynthetase syndrome (ASyS) and immune mediated necrotizing myopathy (IMNM)^1^.

There is a lack of licensed or evidence-based treatments available for patients with IIM, with oral disease modifying therapy largely borrowed from more common rheumatic autoimmune diseases. Glucocorticoids are recommended at high initial doses of 0.5-1mg/kg^2,3^ with variable efficacy, and frequent unwanted adverse effects including steroid myopathy and bone loss. Steroid-sparing drugs are often used off label, e.g. methotrexate, mycophenolate mofetil, tacrolimus, and azathioprine, despite lack of being evidence based for adult patients^2^. Despite a negative phase III trial, rituximab, a B cell depleting antibody targeting CD20, remains a widely used biologic therapy in IIM^4^. Anti-CTLA-4 (abatacept) is used variably despite negative phase III trial data, although a meta-analysis concluded that the drug may improve achievement of efficacy at three or six months^5,6^. Intravenous immunoglobulin is approved by the US Food and Drug Administration for refractory DM^7^ but use remains cost-prohibitive and supply restrictive in many countries. There is thus a high unmet need to identify better investigational medical products for patients with IIM.

Janus Kinases (JAKs) are enzymes that transduce intracellular signals from cell surface receptors for various cytokines and growth factors involved in haematopoiesis, inflammation and immune function. Within the intracellular signalling pathway, JAKs phosphorylate and activate STATs which activate IFN-inducible gene expression within the cell. Type I IFN (a,β) (IFNI) signals via IFNI receptors, to activate TYK2 and JAK1, phosphorylation of STAT1/2, and downstream expression of IFN-stimulated genes. Type II IFN (γ) (IFNII) is dependent on STAT4 phosphorylation, leading to JAK1/JAK2 activation and STAT1 phosphorylation, but can also signal via alternative pathways^8^.

The expression of IFNI-inducible gene transcripts in muscle is high in DM, moderate in ASyS, and low in IMNM and IBM patients^9,10^. Type II IFN inducible genes are highly expressed in DM, IBM, and ASyS, and low in IMNM^10^. Type I IFN-inducible transcripts measured in blood of patients with DM correlate with clinical disease activity measures^11^. Pathogenic *in vitro* effects of type I IFN on muscle have been shown to improve following JAK inhibition^12^.

JAK-STAT inhibition shows promise in IIM, from published case reports and case series with patients demonstrating improvement in mainly skin disease, lung disease and even calcinosis in DM^13^ thus far. A randomized control trial using a TYK2-JAK1 inhibitor, brepocitinib, recently published demonstrated efficacy in patients with DM^14^. We therefore hypothesized that Janus kinase (JAK) pathway plays an active role in the disease process within IIM. We aimed to investigate the clinical efficacy and safety of baricitinib, a selective JAK 1/2 inhibitor, on disease activity in participants with confirmed PM or DM.

## Methods

### Trial design

MYOJAK was a UK, multi-centre, randomized, treatment delayed-start design trial (EudraCT 2019-003868-42) coordinated at the Manchester Clinical Trials Unit and conducted in five sites in the United Kingdom (Bath, London, Leeds, Manchester, Birmingham), in accordance with the Good Clinical Practice guidelines of the International Council for Harmonization and the principles of the Declaration of Helsinki. Research Ethics Committee approval was obtained from UK Health Research Authority (HRA) prior to commencement of the trial (reference 20/NW/0138). Local site permissions were obtained as required in accordance with HRA guidance and institutional requirements. All participants provided informed consent. The trial was monitored by a combined trial steering committee/independent data monitoring committee. The design of the trial was based on the ARTEMIS study, a phase II study of abatacept in IIM^15^ outlined in Figure 1). A copy of the protocol and statistical analysis plan have been uploaded to ClinicalTrials.gov.

**Figure 1:**
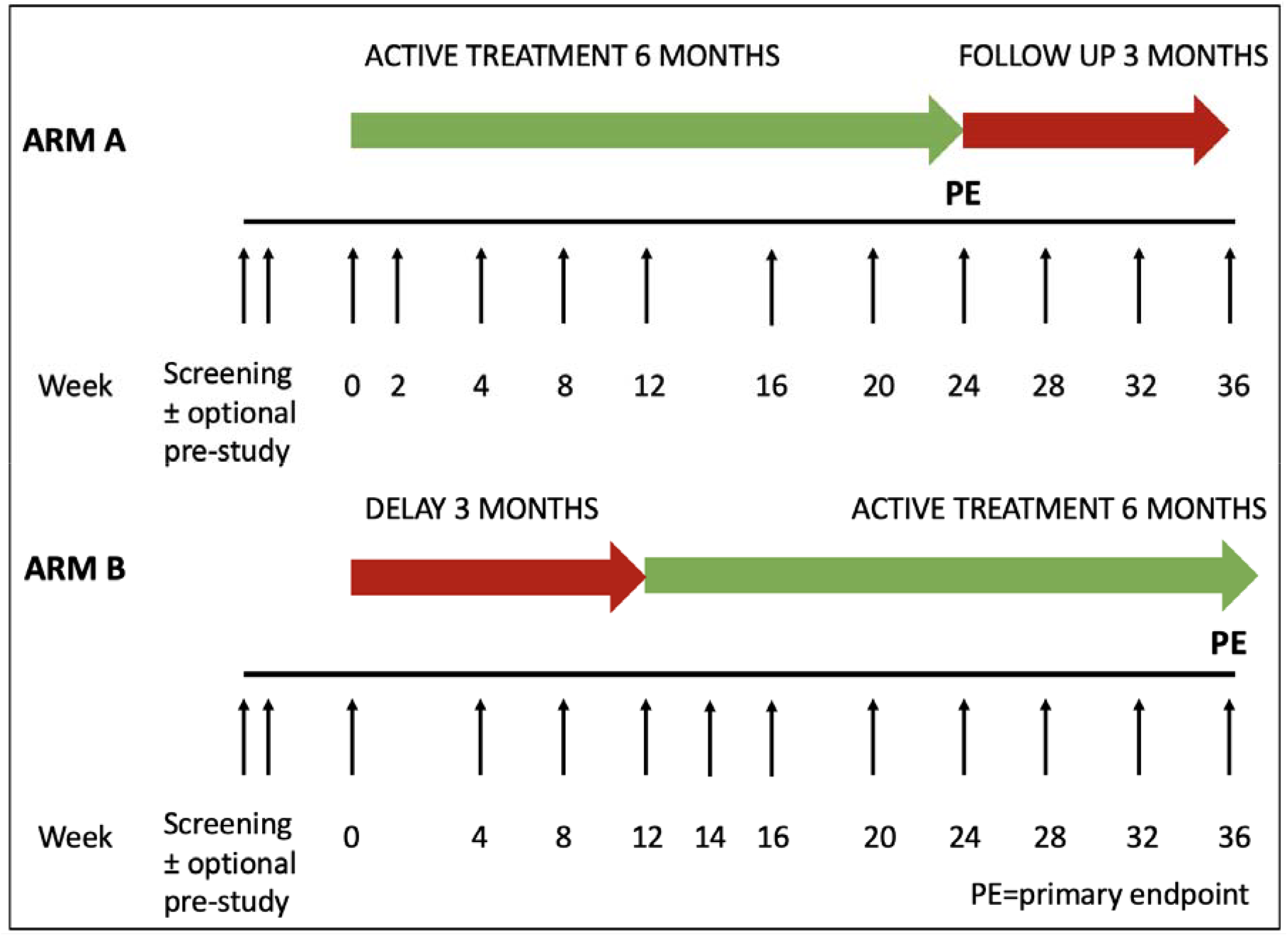
Schedule of assessments

### Patients

Eligible participants were ^3^18 years with a confirmed diagnosis of DM or PM^16^. Patients had persisting disease activity after a minimum of 12 weeks treatment with prednisolone (^3^20mg/day for at least 4 weeks or equivalent). Active inflammatory disease was based on persisting or worsening muscle weakness; bilateral manual muscle testing score (MMT) <150 or low endurance (Functional Index-3 [FI-3] <20% upper value)^17^, and additionally ^3^1 other sign of active disease: i) elevated serum levels of ^3^1 muscle enzyme (CK, LDH, AST, ALT) above upper limit of normal explained by muscle involvement; ii) inflammation on recent muscle biopsy or magnetic resonance imaging (MRI) scans (within 12 weeks); iii) active extramuscular disease (dermatomyositis-specific skin rash, arthritis or interstitial lung disease (ILD) (as suggested by chest x-ray/ high resolution computerised tomography (HRCT) or pulmonary function test (reduction of TLCO by 15% and/or FVC by 10% from baseline)), and on the treating physicians’ judgement. If criteria for active disease were not met, a sum of physician global assessment, patient global assessment and extra muscular global assessment visual analogue scale (VAS) scores ^3^10 cm (all scales individually on 0-10cm scale) was deemed sufficient.

For defined PM, participants were only included if they were positive for myositis-specific (anti-synthetase) or myositis-associated (Ro52, Ro60, PmScl, RNP) autoantibodies, and after being assessed by an adjudication committee formed by myositis experts.

Participants should have received ^3^1 of standard of care medications within the required timeframe: antimalarial (e.g. hydroxychloroquine) for three months at a stable therapeutic dose for at least 8 weeks prior to the screening visit, a single immunosuppressant for three months at a stable therapeutic dose for ^3^4 weeks prior to screening, or an oral glucocorticoid, at a stable dose ≤20 mg/day prednisone (or equivalent), for at least 4 weeks prior to baseline. Full inclusion and exclusion criteria are available in the Supplementary material.

### Patient and Public involvement statement

MYOJAK trial documentation was shared with the University of Manchester Research User Group, an organisation comprised of patient partners, prior to submission to the Ethics Committee. Wording within the documents was adapted and simplified accordingly following feedback.

### Intervention, randomization and blinding

Participants with IIM were screened across the participating centres and following confirmation of eligibility, patients were randomized into one of two arms and received either treatment with 2mg or 4mg baricitinib for 24 weeks, as clinically recommended, from baseline (immediate-start arm), or delayed-start treatment 12 weeks post-randomization (delayed-start arm) (Figure 1). Baricitinib was supploied by Eli Lilly to the participating sites. Participants were equally randomized to either treatment arm using minimisation factors with a computer generated random element controlling for the five sites and confirmed diagnosis of DM or PM. To ensure allocation concealment, the randomization code was not released until the participant had been randomized. As secondary efficacy outcome measures were to be compared between treatment arms, clinician-reported outcome measures were administered and assessed by an efficacy assessor who was blinded to treatment allocation.

### Endpoints/Outcomes

The Total Improvement Score (TIS) is a weighted composite measure of improvement including the six International Myositis Assessment and Clinical Studies Group (IMACS) defined core measures of myositis activity^18^. The primary endpoint was the number of participants achieving at least minimal clinical response (TIS20) as per 2016 American College of Rheumatology/European League Against Rheumatism criteria^19^ 24 weeks post-active treatment (at 24 weeks in the Immediate-start arm, at 36 weeks in the Delayed-start arm). Secondary endpoints included the number of participants achieving at least moderate (TIS40) and major response (TIS60) at 24 weeks post-active treatment, comparison of the clinical response between the treatment arms at 12/24 weeks post-randomization and at 24 weeks post-active treatment, and time taken to achieve TIS20 up to 24 weeks post-active treatment

Further secondary endpoints included change from baseline at 12/24 weeks post-randomization and at 24 weeks post-active treatment between the treatment arms in the following measures: individual components of the IMACS core set measures of disease activity, skin activity assessed by the cutaneous dermatomyositis disease area and severity index (CDASI v2), physical functioning as measured by the Patient Reported Outcomes Measurement Information System (PROMIS PF-20), muscle endurance as tested by the myositis functional index (FI-3), damage assessed by the myositis damage index (MDI), perceived pain measured by a visual analogue scale (VAS), within the Health Assessment Questionnaire, the two body pain related items of The Short Form (36) Health Survey (SF-36) v2, fatigue as measured by the functional assessment of chronic illness therapy (FACIT)-Fatigue questionnaire v4 and components of SF-36v2, health status as measured by the mental and physical sub-scales of the SF-36v2, and health utility as measured by the EQ-5D-5L. The steroid-sparing effect of baricitinib was also assessed across treatment arms after 12 and 24 weeks of active treatment. A tertiary objective included an assessment of the trend in TIS by trial arm over the duration of the trial at baseline, 12, 24, and 36 weeks. Cumulative adverse events (AEs) were assessed across the treatment arms after 12 and 24 weeks of active treatment.

Efficacy was assessed by a physician blinded to treatment arm. A safety assessor, unblinded to the treatment arm, also reviewed the participant, prescribed the medication according to randomization, and assessed and reported on side effects.

### Antibody testing

Myositis antibody testing was conducted at screening. A panel of myositis specific/associated autoantibodies were tested for (Mi-2 alpha, Mi-2 beta, TIF1g, MDA5, NXP2, SAE1, Ku, PM-Scl100, PM-Scl75, Jo-1, SRP, PL-7, PL-12, EJ, OJ, Ro-52, cN-1A) at the University of Bath using a commercially available line immunoassay (Euroimmun, Lübeck, Germany), and ELISA where available (anti-synthetase, MDA5, Jo1, Mi2, TIF1g [MBL], NXP2 [in-house] Ro52 [Organtec]). Local historical antibody results were also available and final autoantibody status was determined by integration of results from all assays.

### Muscle biopsy, MRI imaging, biomarker assessment

An optional muscle biopsy sub-study was carried out to determine whether treatment with baricitinib reduced IFN signalling, type I IFN induced gene expression and markers of cell injury in comparison to usual treatment. Two muscle biopsies were collected for research purposes; one at pre-treatment (between week −2 to 0 for the immediate-start arm and week 8 to 12 for the delayed-start arm) and one at 24 weeks after the start of active treatment (week 24 for the immediate-start arm and week 36 for the delayed-start arm). In participants who consented to the MRI scanning sub-study, two MRI scans were conducted, one at pre-treatment, and one at 12 weeks post-active treatment. Blood and urine samples were collected as part of the main study for future analysis of biomarkers. The results of these assessments will be published separately.

### Statistical analysis

The original intended sample size was 25 patients with outcome data from 20 patients which allows the estimation of achieving at least a minimal response rate of 50% with a two-sided 95% confidence interval (CI) of width no more than 0.456 (i.e. 0.272 to 0.728). However, as the intended sample size was not achieved, the sample size calculation was modified, as suggested by the TSC/IDMC Statistician, and included in the Statistical Analysis Plan. The modified sample size calculation had a two-sided 95% CI of width no more than 0.521 (i.e. 0.266 to 0.787). Baseline characteristics were stratified by the group that participants were randomized to and presented as frequency and percentages for categorical variables and mean/standard deviation for variables with a normal distribution. Treatment effects were estimated by the difference in response rates between treatment arms and 95% exact (binomial) CIs for the differences. Cumulative AEs and glucocorticoid dosing are presented using frequencies and percentages (%) across treatment arms. Primary and secondary efficacy analyses were performed using an intention to treat principle, and the safety population, defined as all participants who received at least one dose of study drug, was used for all safety analyses. A Statistical Analysis Plan was written and signed off prior to the analysis being undertaken. The Trial Protocol and Statistical Analysis Plan can be accessed via clinicaltrials.gov (https://clinicaltrials.gov/study/NCT04208464#study-plan). De-identified participant data can be accessed on request.

## Results

### Recruitment

The MYOJAK trial was conducted between January 2022 and September 2023. The trial was affected by external factors including the Covid-19 pandemic which delayed commencement, and the Manchester Clinical Trials Unit announcing their closure whilst the trial was recruiting which affected recruitment timelines. As illustrated in the CONSORT diagram (Figure 2), 18 patients were recorded in the screening logs as having been screened for eligibility. Two patients were initially screened and were ineligible for the trial due to an ongoing infection and, secondly, being under investigation for other medical reasons. They were re-screened a few weeks later and found to be eligible. In total, three patients were excluded following screening due to being ineligible and declining to participate. Therefore, 15 patients were randomized; 7 allocated to Immediate-Start arm and 8 allocated to Delayed-start arm.

**Figure 2:**
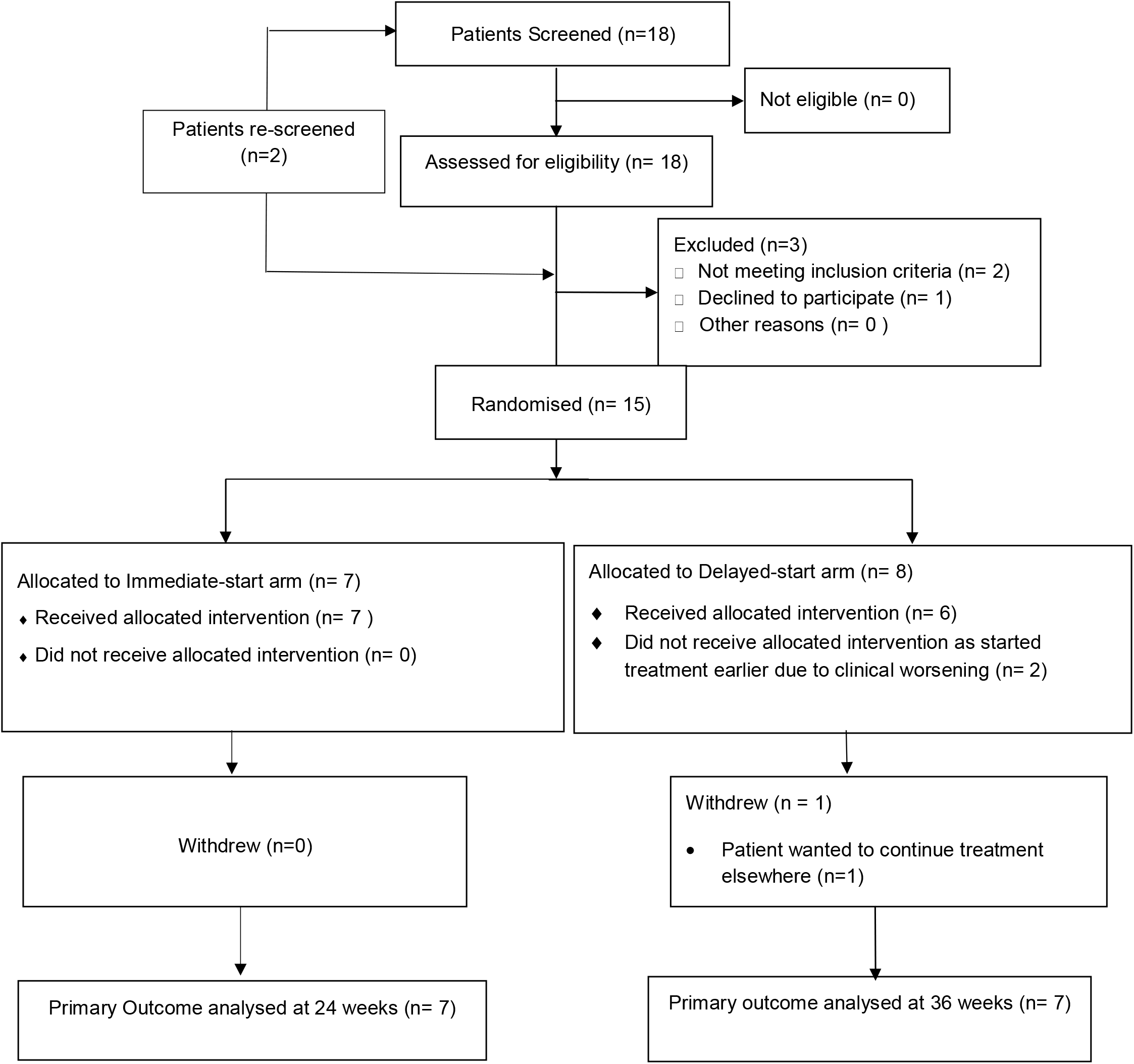
MYOJAK CONSORT Diagram

The original sample size calculation was based on 25 patients with the aim to obtain outcome data from 20 patients. We managed to obtain primary outcome data from 14 out of 15 patients but secondary outcome data up to 24 weeks was available for all 15 patients. The under recruitment is likely to lead to wider confidence intervals and more instability in the estimates.

Two patients randomized to the Delayed-start arm were advised to commence Baricitinib immediately due to clinical worsening (allowed per protocol) and therefore started 24 weeks of treatment at week 0 but were followed up as if they were in the Delayed-start arm up to 36 weeks post-randomization. One patient from the Delayed-start arm withdrew from the study 9 days before the primary outcome visit at 36 weeks post-randomization. Therefore, primary outcome data is only available for 14 out of 15 patients.

### Baseline characteristics

The demographic and clinical characteristics of participants at baseline are summarized in Table 1. The mean (SD) age of patients recruited was 43.2 (11.6) years. More females than males were recruited as one would expect in an IIM population and the majority were White. The confirmed sub-classification for 13 of the 15 patients was DM, and the remaining two were classified with polymyositis (clinical diagnosis of ASyS, both anti-Jo1 positive). Three of 15 (20%) of the patients were seronegative. The majority of patients were taking immunosuppressants at baseline, with two on combination therapy with hydroxychloroquine. Nine out of 12 DM patients had a defined myositis autoantibody.

**Table 1.**
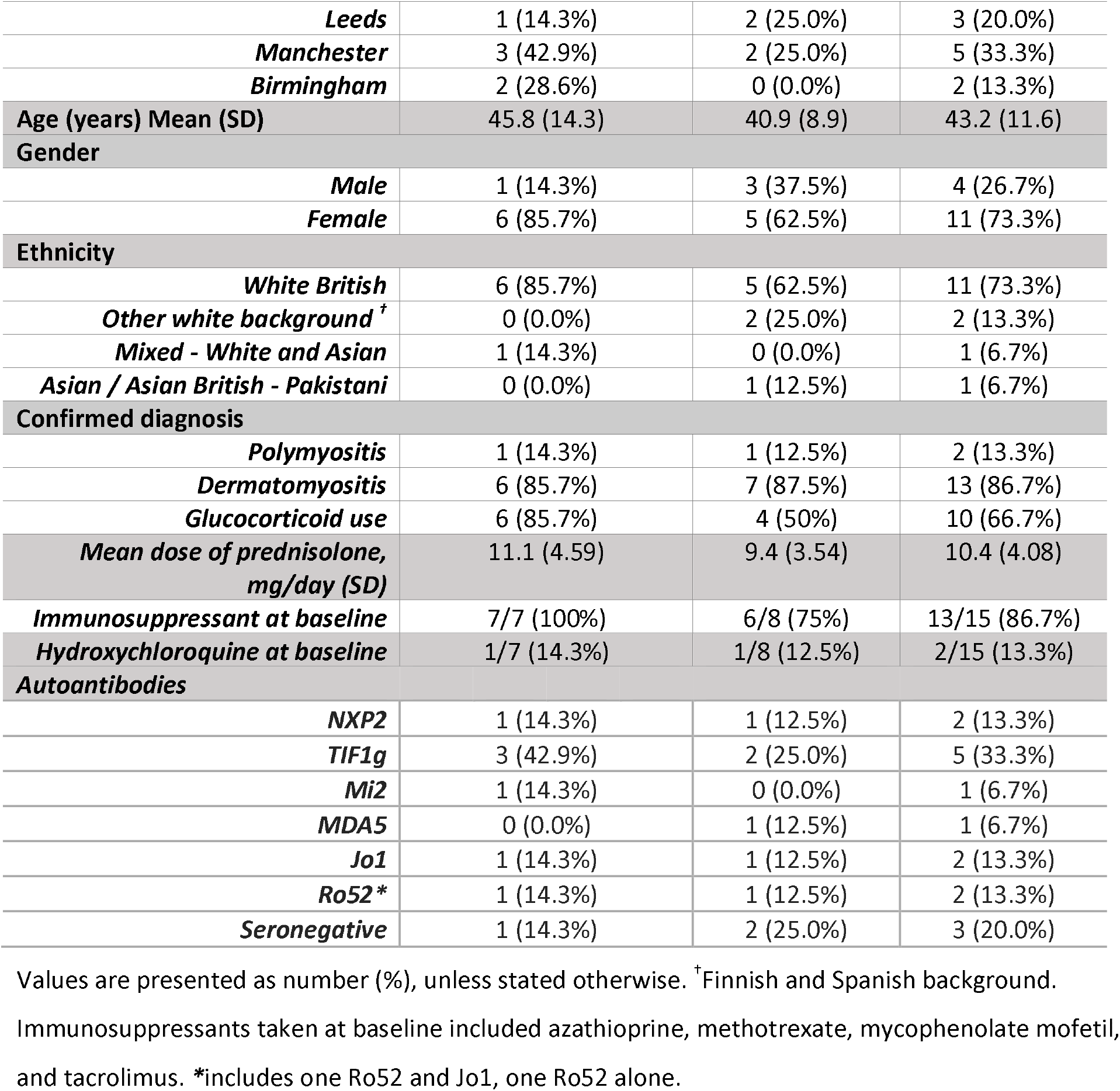
Demographic and clinical characteristics at baseline.

|  |  |  |  |
| --- | --- | --- | --- |
| **Leeds** | 1 (14.3%) | 2 (25.0%) | 3 (20.0%) |
| **Manchester** | 3 (42.9%) | 2 (25.0%) | 5 (33.3%) |
| **Birmingham** | 2 (28.6%) | 0 (0.0%) | 2 (13.3%) |
| **Age (years) Mean (SD)** | 45.8 (14.3) | 40.9 (8.9) | 43.2 (11.6) |
| **Gender** |  |  |  |
| **Male** | 1 (14.3%) | 3 (37.5%) | 4 (26.7%) |
| **Female** | 6 (85.7%) | 5 (62.5%) | 11 (73.3%) |
| **Ethnicity** |  |  |  |
| **White British** | 6 (85.7%) | 5 (62.5%) | 11 (73.3%) |
| **Other white background <sup>†</sup>** | 0 (0.0%) | 2 (25.0%) | 2 (13.3%) |
| **Mixed - White and Asian** | 1 (14.3%) | 0 (0.0%) | 1 (6.7%) |
| **Asian / Asian British - Pakistani** | 0 (0.0%) | 1 (12.5%) | 1 (6.7%) |
| **Confirmed diagnosis** |  |  |  |
| **Polymyositis** | 1 (14.3%) | 1 (12.5%) | 2 (13.3%) |
| **Dermatomyositis** | 6 (85.7%) | 7 (87.5%) | 13 (86.7%) |
| **Glucocorticoid use** | 6 (85.7%) | 4 (50%) | 10 (66.7%) |
| **Mean dose of prednisolone, mg/day (SD)** | 11.1 (4.59) | 9.4 (3.54) | 10.4 (4.08) |
| **Immunosuppressant at baseline** | 7/7 (100%) | 6/8 (75%) | 13/15 (86.7%) |
| **Hydroxychloroquine at baseline** | 1/7 (14.3%) | 1/8 (12.5%) | 2/15 (13.3%) |
| **Autoantibodies** |  |  |  |
| **NXP2** | 1 (14.3%) | 1 (12.5%) | 2 (13.3%) |
| **TIF1g** | 3 (42.9%) | 2 (25.0%) | 5 (33.3%) |
| **Mi2** | 1 (14.3%) | 0 (0.0%) | 1 (6.7%) |
| **MDA5** | 0 (0.0%) | 1 (12.5%) | 1 (6.7%) |
| **Jo1** | 1 (14.3%) | 1 (12.5%) | 2 (13.3%) |
| **Ro52\*** | 1 (14.3%) | 1 (12.5%) | 2 (13.3%) |
| **Seronegative** | 1 (14.3%) | 2 (25.0%) | 3 (20.0%) |

Values are presented as number (%), unless stated otherwise. <sup>†</sup>Finnish and Spanish background.
Immunosuppressants taken at baseline included azathioprine, methotrexate, mycophenolate mofetil, and tacrolimus. \*includes one Ro52 and Jo1, one Ro52 alone.

### Baseline disease activity

The baseline disease activity measures of the participants are summarized in Table 2. No evidence of a difference were noted between the Delayed-start and the Immediate-start arm. The trial participants exhibited both active skin (CDASI-A 20.8 [11.1]) as well as muscle (bilateral MMT8, 131.3 [18]) disease. The HAQ disability index demonstrated a low to moderate level of disability at baseline. The level of skeletal muscle enzymes is summarized in Supplementary Table 1, which were numerically higher in the Delayed-start compared to the Immediate-start arm, influenced by one participant with high baseline enzyme levels. Patients had minimal disease damage according to the myositis damage index (MDI) (extent 0.1 [0.1], severity 0.6 [0.3].

**Table 2.** Disease activity measures at baseline.

|  | Immediate-Start (N = 7) | Delayed-Start (N = 8) | Total (N = 15) |
| --- | --- | --- | --- |
| **Physician global assessment of disease activity VAS (0-10)** | 4.5 (2.1) | 5.7 (2.4) | 5.1 (2.2) |
| **MMT8 score (0-150)** | 133.7 (13.4) | 129.1 (22.0) | 131.3 (18.0) |
| **Patient global assessment of disease activity VAS (0-100)** | 6.2 (1.8) | 7.4 (1.4) | 6.8 (1.7) |
| **MDAAT tool (0-10)** | 1.3 (0.4) | 1.4 (0.7) | 1.3 (0.6) |
| **HAQ (0-3)** | 1.4 (0.5) | 1.4 (1.0) | 1.4 (0.8) |
| **CDASiv2** |  |  |  |
| **CDASI Total Activity score (0-99)** | 21.8 (5.6) | 19.9 (14.8) | 20.8 (11.1) |
| **CDASI Total Damage score (0-32)** | 5.2 (4.1) | 3.7 (6.6) | 4.4 (5.4) |
| **FI-3 score (0-150)** | 17.0 (7.5) | 18.6 (14.8) | 17.8 (11.6) |
| **PROMIS PF-20 T-score (9.2 – 62.7)** | 34.9 (5.5) | 38.5 (14.1) | 36.8 (10.7) |
| **EQ-5D-5L utility** | 0.5 (0.2) | 0.6 (0.3) | 0.6 (0.3) |
| **SF-36 MCS score** | 41.5 (7.0) | 46.9 (8.0) | 44.6 (7.8) |
| **SF-36 PCS score** | 35.1 (9.7) | 35.6 (12.7) | 35.4 (11.1) |
| **Perceived Pain VAS (0-100)** | 46.1 (19.0) | 40.1 (27.7) | 43.1 (23.0) |
| **FACIT-Fatigue (0-52)** | 30.1 (10.1) | 28.2 (11.8) | 29.1 (10.7) |

Note: Mean (SD) presented. Key: MMT = manual muscle testing, VAS = visual analogue scale, MDAAT = Myositis Disease Activity Assessment Tool, HAQ = health assessment questionnaire, CDASI = Cutaneous Dermatomyositis Disease Area and Severity Index, FI-3 = functional index-3, PROMIS PF-20 = Patient-Reported Outcomes Measurement Information System - Physical Function 20-item short form, SF-36 = Short-Form Health Survey, MCS = Mental Component Summary, PCS = Physical Component Summary, FACIT = Functional Assessment of Chronic Illness Therapy.

### Primary outcome

The primary outcome of at least minimal clinical response according to the IMACS criteria, TIS20, at 24 weeks post-active treatment (at 24 weeks in the Immediate-start arm and at 36 weeks in the Delayed-start arm) was achieved in 14 of 15 (93%) patients (Table 3). One patient withdrew from the trial prior to the primary outcome data being collected. The patient attended an early termination visit 9 days prior to the end trial visit. From information based on the early termination visit data, this patient also achieved minimal clinical response.

**Table 3:**
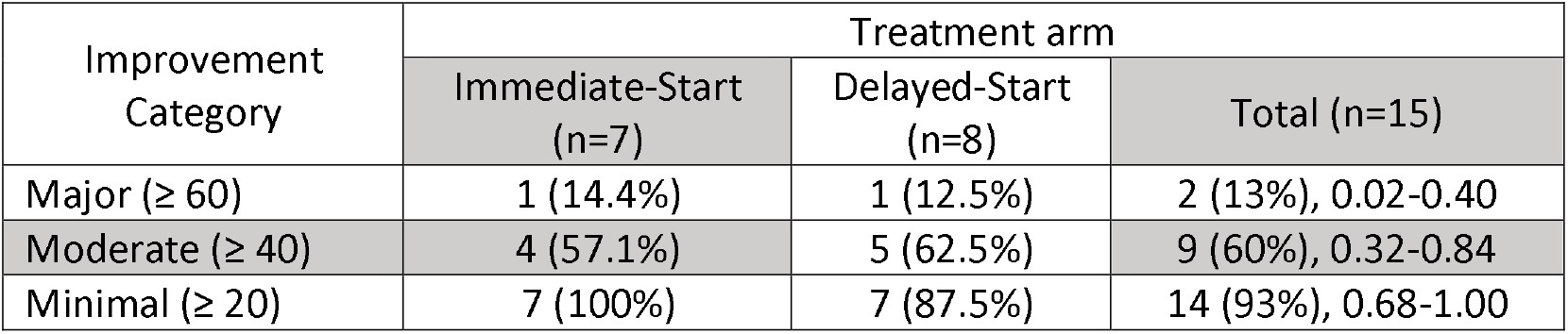
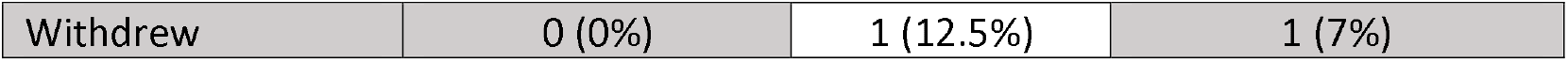
Improvement Category at 24 weeks post-active treatment with baricitinib.

### Secondary outcomes

#### Extent of clinical response across treatment arms after 24 weeks of active treatment

Nine out of 15 (60%) achieved at least moderate clinical response, TIS40, and two out of 15 (13%) achieved at least major clinical response, TIS60 (Table 3). At 24 weeks post-treatment, there was little evidence of a difference in response rates for those patients who achieve at least moderate clinical response in the Immediate- or Delayed-start arms (−0.05, 95% CI −0.55-0.44).

#### Comparison of clinical response between treatment arms at 12/24 weeks post-randomization

The number of patients who achieved TIS20 at 12 weeks post-randomization was investigated to assess the effect of commencement of treatment in the Immediate-start arm versus those on standard treatment (Immediate-start arm patients who received 12 weeks of treatment, Delayed-start patients yet to start treatment). Eleven out of 15 (73%) patients achieved at least TIS20 (95% CI 0.45-0.92), including all Immediate-start arm patients and four Delayed-start arm patients.

Fourteen out of 15 (93%) achieved at least TIS20 (95% CI 0.68-1.00) at 24 weeks post-randomization (Immediate-start arm patients who have completed 24 weeks treatment and Delayed-start arm patients who completed 12 weeks). Any differences between the arms were lost at 24 weeks post-randomization by the time Delayed-start arm patients had received baricitinib for 12 weeks.

On comparing the Immediate- and Delayed-start arm data at 12 weeks post-randomization (Immediate-start arm patients who have completed 12 weeks treatment and Delayed-start arm patients yet to start baricitinib) (Table 4), evidence of a difference was noted for two of the IMACS core set measures, the patient global activity score, and MDAAT. Evidence of improvement in skin was noted as assessed by the CDASI activity score. Finally, evidence of improvements in pain (SF-36 bodily pain and HAQ pain VAS), fatigue (FACIT-Fatigue and Fatigue on SF-36), and SF-36 mental/physical health scores were also noted.

**Table 4:** Change at 12 weeks post-randomization.

| Measure | Treatment arm |  | Mean difference (95% CI) |
| --- | --- | --- | --- |
|  | Immediate-Start<br>(N = 7) | Delayed-Start (N = 8) |  |
| **Physician global assessment of disease activity VAS (0-10)** | -2.27 (1.8) | -0.70 (2.2) | -1.6 (-3.5 to 0.4) |
| **Patient global assessment of disease activity VAS (0-10)** | -2.95 (1.7) | -0.04 (1.4) | -2.9 (-4.4 to -1.4) |
| **MMT-8** | 11.57 (9.6) | 5.25 (11.6) | 6.3 (-4.1 to 16.7) |
| **HAQ<sup>1</sup>** | -0.08 (0.4) | 0.05 (0.3) | -0.1 (-0.5 to 0.3) |
| **Muscle Enzyme: CK<sup>2</sup>** | -37.14 (164.06) | -307.57 (822.1) | 270.4 (-310.8 to 851.6) |
| **Muscle Enzyme: AST or ALT** | 1.29 (11.7) | -13.38 (42.9) | 14.7 (-15.7 to 45.0) |
| **Muscle Enzyme: LDH** | 17.14 (72.5) | -27.25 (81.3) | 44.4 (-33.1 to 121.8) |
| **MDAAT** | -0.73 (0.3) | 0.31 (0.8) | -1.0 (-1.6 to -0.5) |
| **CDASI Total Activity Score<sup>3</sup>** | -8.67 (4.9) | -1.43 (5.9) | -7.2 (-13.1 to -1.3) |
| **CDASI Total Damage Score<sup>3</sup>** | 0.0 (4.4) | 0.14 (1.2) | -0.14 (-3.8 to 3.5) |
| **PROMIS PF-20 T-score** | 1.83 (3.9) | -2.34 (5.5) | 4.2 (-0.6 to 8.9) |
| **FI-3 score** | 7.29 (9.3) | 1.29 (12.6) | 6.0 (-5.1 to 17.1) |
| **MDI Tool** |  |  |  |
| ***MDI extent of damage score*** | -0.03 (0.1) | 0.03 (0.1) | -0.1 (-0.1 to 0.0) |
| ***MDI severity of damage score*** | -0.08 (0.5) | 0.08 (0.3) | -0.2 (-0.5 to 0.2) |
| ***MDI extended damage score<sup>4</sup>*** | -0.03 (0.1) | 0.01 (0.1) | -0.0 (-0.1 to 0.0) |
| **Perceived Pain** |  |  |  |
| ***Bodily Pain (SF-36)*** | 23.43 (8.44) | -8.75 (19.0) | 32.2 (17.8 to 46.6) |
| ***HAQ VAS<sup>2</sup>*** | -15.32 (17.0) | -4.1 (19.1) | -14.9 (-33.5 to 3.7) |
| **Fatigue** |  |  |  |
| ***FACIT-Fatigue*** | 8.57 (3.4) | -4.50 (6.4) | 13.1 (8.1 to 18.1) |
| ***Fatigue (SF-36)*** | 24.11 (14.6) | 1.56 (14.1) | 22.5 (8.5 to 36.5) |
| **Health Status** |  |  |  |
| ***SF-36 MCS<sup>3</sup>*** | 7.79 (4.9) | 2.37 (3.0) | 5.4 (1.1 to 9.8) |
| ***SF-36 PCS<sup>3</sup>*** | 6.03 (4.1) | -1.55 (4.0) | 7.6 (3.3 to 11.9) |
| ***EQ-5D-5L Health Utility<sup>5</sup>*** | 0.1 (0.2) | -0.1 (0.2) | 0.2 (0.0 to 0.4) |

Values are presented as mean (SD).<sup>1</sup> One patient did not have the measure assessed at 12 weeks. <sup>2</sup>One patient did not have the measure assessed at baseline. <sup>3</sup>Two patients have missing data, one in each Treatment group. <sup>4</sup>One patient in the Immediate-Start group had missing data therefore unable calculate the MDI extended damage score. <sup>5</sup>One patient in the Delayed-Start group did not have the measure assessed at week 12. Effect estimate was calculated as: mean difference = Immediate-start – Delayed-start. Key: VAS = visual analogue scale, MMT = manual muscle testing, HAQ = health assessment questionnaire, CK = creatine kinase, AST = aspartate aminotransferase, ALT = alanine aminotransferase, LDH = lactate dehydrogenase, MDAAT = Myositis Disease Activity Assessment Tool, CDASI = Cutaneous Dermatomyositis Disease Area and Severity Index, PROMIS PF-20 = Patient-Reported Outcomes Measurement Information System - Physical Function 20-item short form, FI-3 = functional index-3, MDI = myositis damage index, SF-36 = Short-Form Health Survey, FACIT = Functional Assessment of Chronic Illness Therapy, MCS = Mental Component Summary, PCS = Physical Component Summary.

Numerical improvements were also noted for physical function as evidenced by the HAQ score, which exceeded the accepted minimally clinically important difference within the Immediate-start arm^19^. Whilst an improvement was noted for the PROMIS PF-20 score in the Immediate-start arm, a slight reduction was noted in the Delayed-start arm.

The differences noted between the Immediate- and Delayed-start arms largely disappeared at 24 weeks post-randomization after the Delayed-start arm patients had received 12 weeks of active treatment (Supplementary table 2).

#### Change from baseline in individual outcome measures

Table 5 shows the change in clinical parameters from baseline at 24 weeks post-active treatment. No evidence of differences were noted between the Delayed-start and Immediate-start arm. Improvements were noted for muscle disease, as evidenced by the change in MMT8 and muscle enzymes, as well as an improvement in the functional index-3 score. No deterioration in the myositis damage index was noted. An improvement in skin disease was also noted as evidenced by a reduction of the CDASI score in both treatment arms.

**Table 5:**
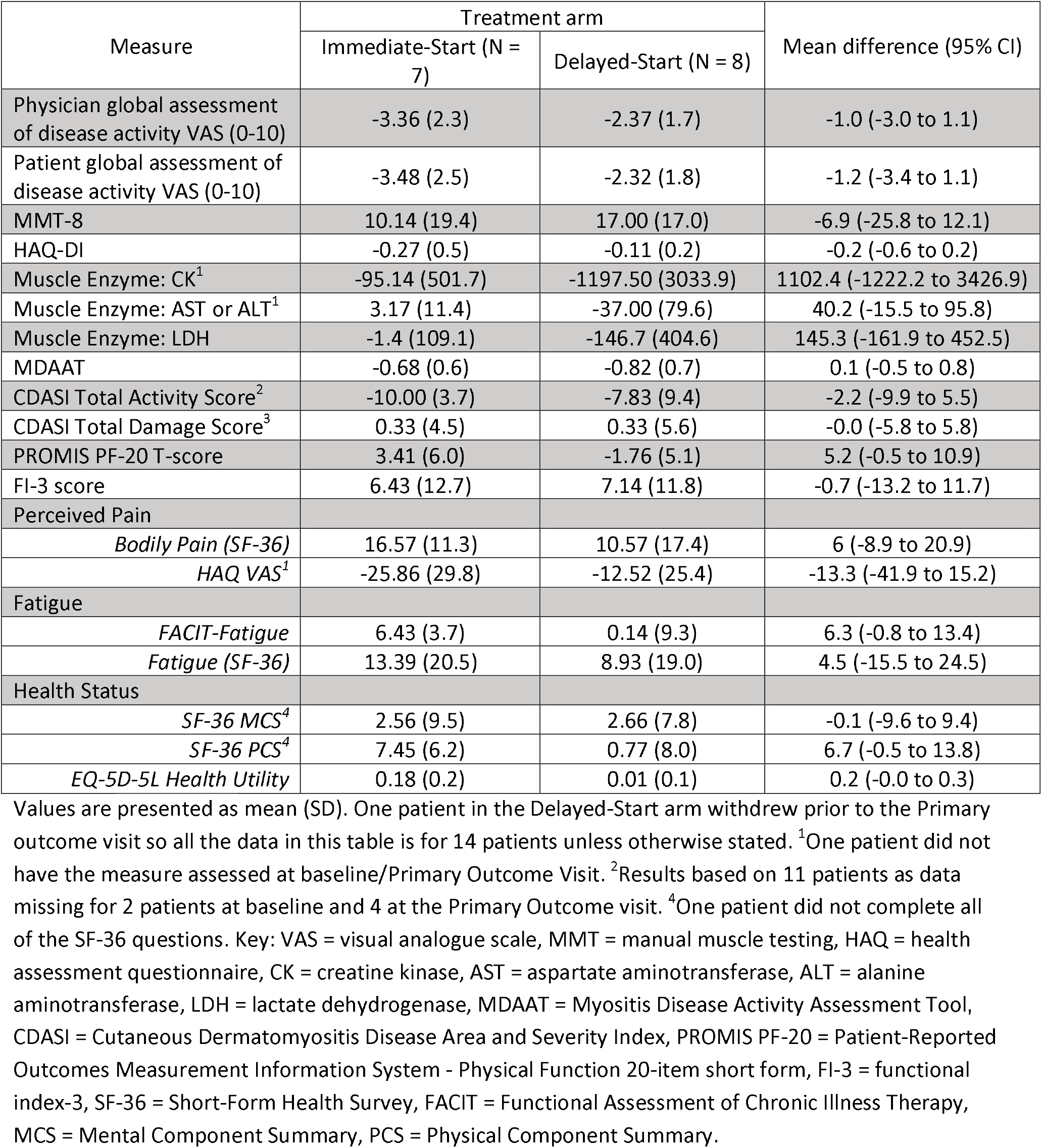
Change from baseline to 24-weeks post-active treatment.

| Measure | Treatment arm |  | Mean difference (95% CI) |
| --- | --- | --- | --- |
|  | Immediate-Start (N = 7) | Delayed-Start (N = 8) |  |
| Physician global assessment of disease activity VAS (0-10) | -3.36 (2.3) | -2.37 (1.7) | -1.0 (-3.0 to 1.1) |
| Patient global assessment of disease activity VAS (0-10) | -3.48 (2.5) | -2.32 (1.8) | -1.2 (-3.4 to 1.1) |
| MMT-8 | 10.14 (19.4) | 17.00 (17.0) | -6.9 (-25.8 to 12.1) |
| HAQ-DI | -0.27 (0.5) | -0.11 (0.2) | -0.2 (-0.6 to 0.2) |
| Muscle Enzyme: CK<sup>1</sup> | -95.14 (501.7) | -1197.50 (3033.9) | 1102.4 (-1222.2 to 3426.9) |
| Muscle Enzyme: AST or ALT<sup>1</sup> | 3.17 (11.4) | -37.00 (79.6) | 40.2 (-15.5 to 95.8) |
| Muscle Enzyme: LDH | -1.4 (109.1) | -146.7 (404.6) | 145.3 (-161.9 to 452.5) |
| MDAAT | -0.68 (0.6) | -0.82 (0.7) | 0.1 (-0.5 to 0.8) |
| CDASI Total Activity Score<sup>2</sup> | -10.00 (3.7) | -7.83 (9.4) | -2.2 (-9.9 to 5.5) |
| CDASI Total Damage Score<sup>3</sup> | 0.33 (4.5) | 0.33 (5.6) | -0.0 (-5.8 to 5.8) |
| PROMIS PF-20 T-score | 3.41 (6.0) | -1.76 (5.1) | 5.2 (-0.5 to 10.9) |
| FI-3 score | 6.43 (12.7) | 7.14 (11.8) | -0.7 (-13.2 to 11.7) |
| Perceived Pain |  |  |  |
| *Bodily Pain (SF-36)* | 16.57 (11.3) | 10.57 (17.4) | 6 (-8.9 to 20.9) |
| *HAQ VAS<sup>1</sup>* | -25.86 (29.8) | -12.52 (25.4) | -13.3 (-41.9 to 15.2) |
| Fatigue |  |  |  |
| *FACIT-Fatigue* | 6.43 (3.7) | 0.14 (9.3) | 6.3 (-0.8 to 13.4) |
| *Fatigue (SF-36)* | 13.39 (20.5) | 8.93 (19.0) | 4.5 (-15.5 to 24.5) |
| Health Status |  |  |  |
| *SF-36 MCS<sup>4</sup>* | 2.56 (9.5) | 2.66 (7.8) | -0.1 (-9.6 to 9.4) |
| *SF-36 PCS<sup>4</sup>* | 7.45 (6.2) | 0.77 (8.0) | 6.7 (-0.5 to 13.8) |
| *EQ-5D-5L Health Utility* | 0.18 (0.2) | 0.01 (0.1) | 0.2 (-0.0 to 0.3) |

Values are presented as mean (SD). One patient in the Delayed-Start arm withdrew prior to the Primary outcome visit so all the data in this table is for 14 patients unless otherwise stated. <sup>1</sup>One patient did not have the measure assessed at baseline/Primary Outcome Visit. <sup>2</sup>Results based on 11 patients as data missing for 2 patients at baseline and 4 at the Primary Outcome visit. <sup>4</sup>One patient did not complete all of the SF-36 questions. Key: VAS = visual analogue scale, MMT = manual muscle testing, HAQ = health assessment questionnaire, CK = creatine kinase, AST = aspartate aminotransferase, ALT = alanine aminotransferase, LDH = lactate dehydrogenase, MDAAT = Myositis Disease Activity Assessment Tool, CDASI = Cutaneous Dermatomyositis Disease Area and Severity Index, PROMIS PF-20 = Patient-Reported Outcomes Measurement Information System - Physical Function 20-item short form, FI-3 = functional index-3, SF-36 = Short-Form Health Survey, FACIT = Functional Assessment of Chronic Illness Therapy, MCS = Mental Component Summary, PCS = Physical Component Summary.

#### Time taken to achieve TIS20 across treatment arms

In the Immediate-start arm, TIS20 was achieved as early as four weeks post-randomization (Figure 3). In the Delayed-start arm, TIS20 was achieved by 24 weeks post-randomization (12 weeks post-active treatment).

**Figure 3:**
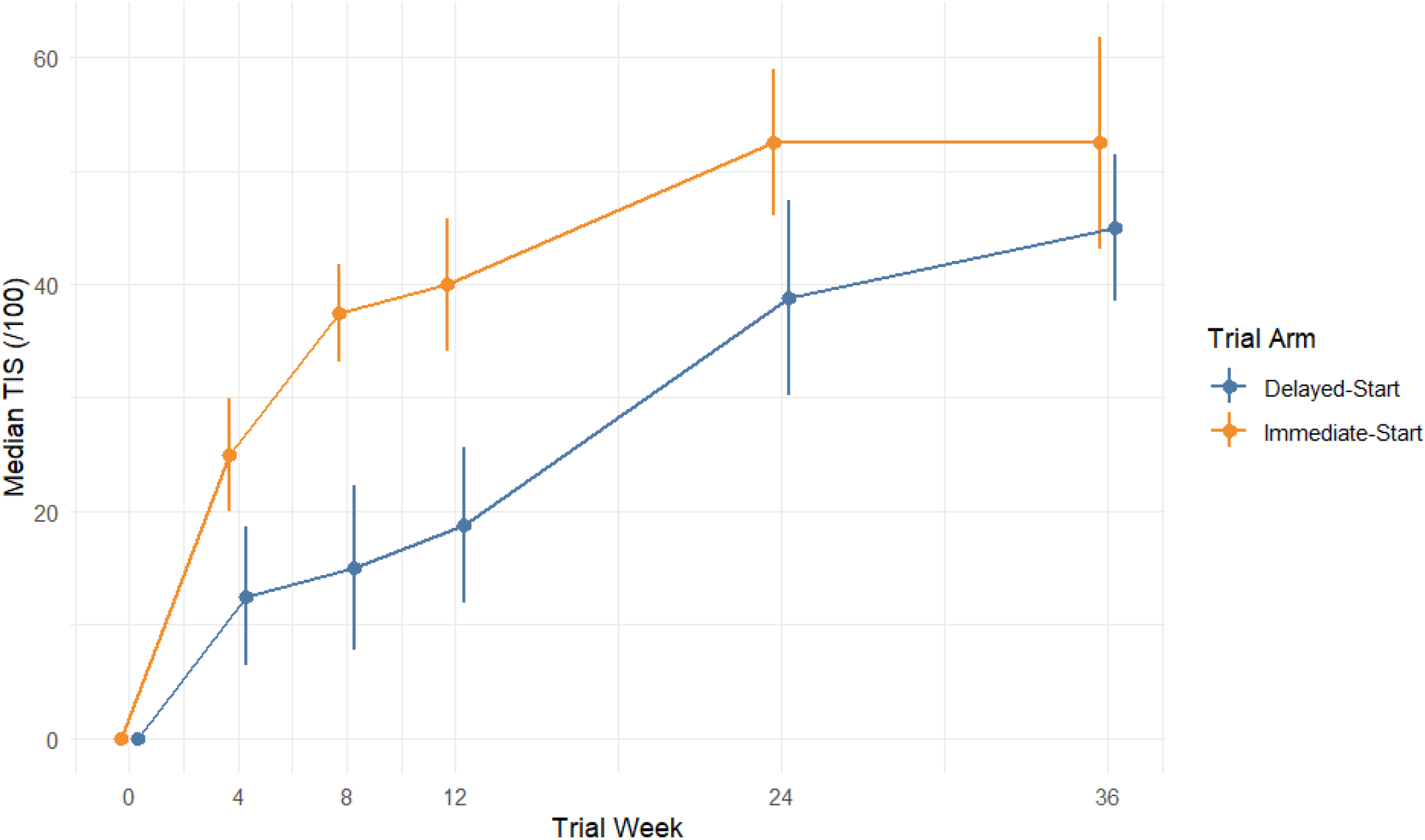
Longitudinal change in Total Improvement Score (TIS) from baseline Median TIS reported alongside error bars that denote interquartile ranges. Immediate-start arm received active Baricitinib treatment from Week 0-24 and delayed-start arm received active Baricitinib treatment from Week 12-36. Immediate-start arm n=7. Delayed-start arm n=8.

#### Time taken to achieve TIS40 at 24 weeks post-active treatment

Figure 3 shows the median TIS plotted over the duration of the trial, comparing the Immediate-to the Delayed-start arm. The Immediate-start arm achieved TIS40 by week 12 but by week 24 the TIS response overlapped for both treatment groups.

#### Glucocorticoid use

At 24 weeks post-active treatment, the majority of patients were still taking glucocorticoids (Immediate-start: 85.7%, average daily dose 9.0mg [3.47]; Delayed-start: 62.5%, 7.1mg [2.13]). The suggested taper regime outlined in the protocol was not mandatory to undertake over the trial duration.

#### Adverse events

There were a total of 58 adverse events (AEs) during the trial (Supplementary Table 3). By severity, there were 34 classed as mild severity, 20 classed as moderate severity and 4 classed as severe severity. The severe AEs included a laceration in R shin following a fall, urinary tract infection, aseptic necrosis in femur heads (bilateral), and drug reaction to Cyclophosphamide. There was one serious AE (SAE) in each treatment arm with one due to a vascular disorder (SAE grade 2) and the other due to skin and subcutaneous disease (SAE grade 3) both requiring hospitalization but not regarded as related to the trial drug.

## Discussion

In this phase IIa treatment delayed-start design trial of an oral JAK 1/2 inhibitor baricitinib for adult patients with IIM, the prespecified primary outcome of TIS20 at 24 weeks post-active treatment was met by 93% of the participating patients. Furthermore, 60% of patients met the secondary outcome of TIS40. A rapid onset of action was also demonstrated with the majority of patients responding to active treatment within 12 weeks. Within the Immediate start arm, a TIS20 response was noted as early as 4 weeks, and a TIS40 at 12 weeks. The Delayed-start design of the trial allowed an analysis between those patients already taking baricitinib at 12 weeks with those still on background treatment, demonstrating a significant divergence in TIS at 12 weeks post-randomization, although caution should be applied to interpretation due to the limited number of recruited participants. At 12 weeks post-randomization, there is evidence of differences between the Immediate-start and Delayed-start arms for patient global, extramuscular, CDASI skin activity, pain, fatigue and SF-36 mental/physical health scores. At baseline, the majority of enrolled participants were classified as having DM, had evidence of both active skin and muscle disease and were treatment experienced. Notable improvements were noted for both the skin and muscle components of disease. Improvements were also noted in endurance, pain, quality of life and fatigue scores.

The positive results in our trial, where the predominant clinical subgroup was DM, are well in line with reports showing beneficial effects of JAK inhibitors in patients with DM, including an open trial using tofacitinib^20^, and in the recently published randomized controlled trial using brepocitinib, a TYK2/JAK1 inhibitor, over 52 weeks^14^. In the brepocitinib trial 68% of the patients reached TIS40 compared with 44% in the placebo arm at 52 weeks. In line with previous reports using JAK inhibitors we also observed an improvement in skin rash as assessed by CDASI.

Baricitinib was overall well tolerated in both treatment arms, demonstrating good safety and tolerability with no unexpected events noted. The trial had already started before regulatory guidance became apparent regarding events of special interest for JAK inhibitors, including venous thromboembolic events, increased risk of cardiovascular disease and cancer. Nevertheless, no such events were noted during the study. As such events appear to be prevalent in myositis spectrum disease, it will be important to monitor patients with IIM on long-term JAK inhibition.

A number of limitations should be noted. The majority of patients did achieve TIS20, including those not on active treatment. At the time of trial design, the TIS criteria had not been properly tested in a clinical trial setting. A high placebo response for TIS20 has since been noted^14,21^. Whilst steroid-sparing has been demonstrated in a further JAK inhibitor study, the MYOJAK trial was not powered to demonstrate this finding, and furthermore there was no mandatory steroid tapering during the study. Due to the delayed-start design there was a lack of placebo within the trial, and only the efficacy assessor was blinded, not the patient. Inadvertent unblinding of efficacy assessors would have been a risk as participants will have been aware of the timing of their active treatment. Measures were taken to prevent unblinding, e.g. by clearly identifying and concealing trial documents containing unblinded information from the efficacy assessor, and reminding participants prior to each trial visit not to disclose their arm allocation. Further long-term efficacy data will be required to establish the sustainability of JAK inhibition in controlling disease.

Prior case series have not established the efficacy within muscle inflammation specifically and therefore the MYOJAK results provide reassurance. Furthermore, improvements were also noted in the two patients with ASyS which has not previously been demonstrated in a clinical trial. The mechanism of type I and type II IFN inhibition suggests that this class of drugs will be beneficial in IIM subtypes, including DM and ASyS. The caveat is that different JAK inhibitors have different modes of action depending on which component of JAK or TYK2 is being inhibited, which may broaden further the mode of action and indications. Other therapeutic strategies for IFN inhibition include targeting the IFN receptor subunit 1 receptor and IFNβ.

Newer B and plasma cell immune strategies targeting CD19, CD20, CD38, BCMA are also in development^9^, and other therapeutic targets including FCRn inhibition are currently under consideration ^22 23^. It remains to be seen in the future which sub-types of IIM are best targeted by which therapeutic target.

In conclusion, the MYOJAK study has demonstrated that baricitinib has a favourable effect in patients with adult IIM after 24 weeks of treatment, in reducing muscle and skin disease activity as well as improving muscle function and quality of life, with no notable safety adverse events. Furthermore, the treatment effect was observed by 12 weeks. However, a randomized placebo-controlled trial is needed to confirm the efficacy of baricitinib in patients with IIM.

## Data Availability

All data produced in the present study are available upon reasonable request to the authors

## Acknowledgements and affiliations

The study has previously been presented in abstract format:

Chinoy H, Krishan A, Sylvestre Y, Lilleker J, Gordon P, Tansley S, Prabu A, Aslam A, Snedden A, Lamb J. Baricitinib in the Treatment of Adult Idiopathic Inflammatory Myopathy: A Randomized, Treatment Delayed-Start Clinical Trial [abstract]. *Arthritis Rheumatol*. 2024; 76 (suppl 9). https://acrabstracts.org/abstract/baricitinib-in-the-treatment-of-adult-idiopathic-inflammatory-myopathy-a-randomized-treatment-delayed-start-clinical-trial/.

The study was supported by an Investigator Initiated Research grant from Eli Lilly and Company and provision of the study drug (Baricitinib). This study has been delivered through the National Institute for Health and Care Research (NIHR) Biomedical Research Centre: Manchester (NIHR203308) and carried out at the NIHR Manchester Clinical Research Facility (CRF) (NIHR203956). The views expressed are those of the author(s) and not necessarily those of the NIHR or the Department of Health and Social Care. Fionnuala Palmer coordinated the study and all the patient visits at Bath, UK. Manchester Clinical Research Facility staff: Smitha Joseph, Sujamole Subin, Tolga Turgut, Sheilla Achieng.

HC has received grant support from Pfizer; has received consulting fees from argenx, Astra Zeneca, Boehringer Ingelheim, Pfizer, J&J, Xencor; speaking engagements from CabalettaBio, Data and Science Monitoring Board chair for Horizon Therapeutics;

AP has received speaker fees from Boehringer Ingelheim.

JAL has received grant support from Pfizer.

ST has received honorarium from Boehringer Ingelheim, Bamboo medical and Priovant

JBL has received advisory board payments from Roche and Abcuro, speakers fees from Roche and Dyne pharmaceuticals, research support from Roche, travel/conference support from Roche and AstraZeneca.

IEL has funding support from: Swedish Research Council, 2024-02582, the Swedish Rheumatism Association, R-1013188; Region Stockholm RS2022-0674, King Gustaf V 80th Year Foundation FAI-2024-1092.

I.E.L. has received honorarium for lecture from Boehringer Ingelheim, research grant from Astra Zeneca and Janssen Pharmaceutica NV, and has been serving on the advisory board for Astra-Zeneca, argenx, Chugai, Novartis, Pfizer and Janssen Pharmaceutica NV, and has stock shares in Roche and Novartis.

## Key messages

Please summarise the key points of your manuscript in a few bullet points under the following headings:

### What is already known on this topic

There is a lack of licensed or evidence-based treatments available in idiopathic inflammatory myopathy. The JAK/STAT pathway plays a key role in the pathogenesis of idiopathic inflammatory myopathy.

### What this study adds

Baricitinib works for both skin and muscle disease in polymyositis and dermatomyositis and also improves quality of life.

### How this study might affect research, practice or policy

The study provides the potential for repurposing an existing licenced drug in a rare disease indication.

|  | Immediate-Start (N = 7) | Delayed-Start (N = 8) | Overall (N = 15) |
| --- | --- | --- | --- |
| Sites |  |  |  |
| *Bath* | 0 (0.0%) | 1 (12.5%) | 1 (6.7%) |
| *King's College Hospital* | 1 (14.3%) | 3 (37.5%) | 4 (26.7%) |

**Supplementary table 1:**
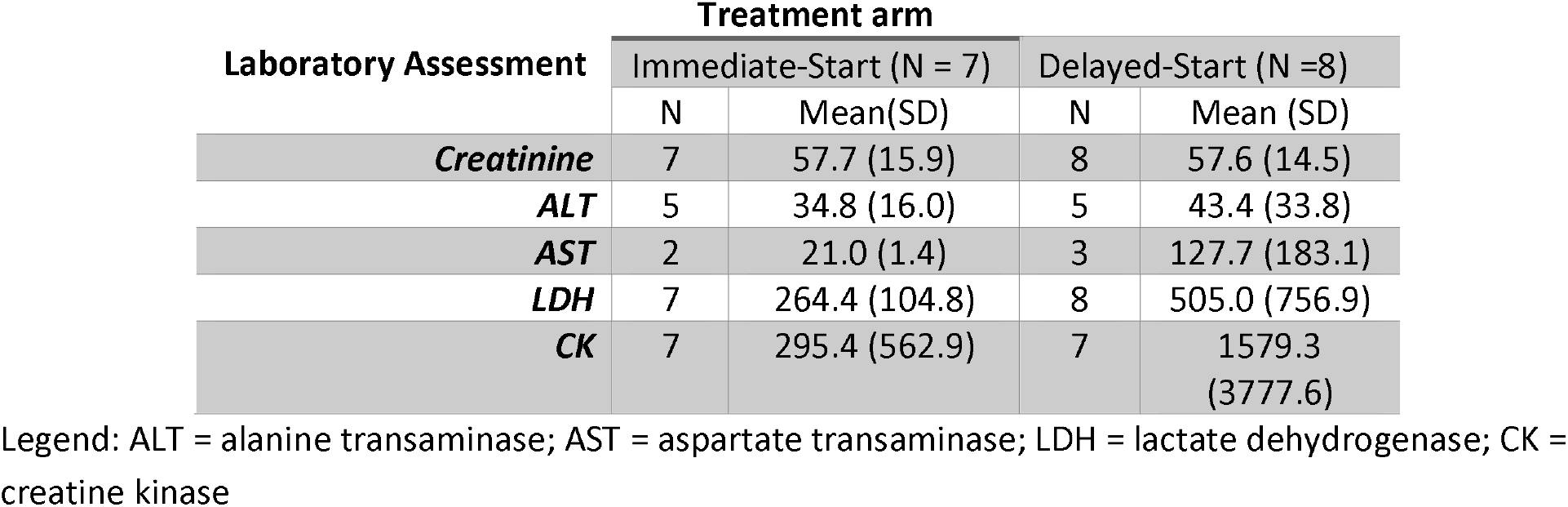
Laboratory Assessments at Baseline.

**Supplementary Table 2:** Change at 24 weeks post-randomization.

| Measure | Treatment arm |  | Mean difference (95% CI) |
| --- | --- | --- | --- |
|  | Immediate-Start (N = 7) | Delayed-Start (N = 8) |  |
| **Physician global assessment of disease activity VAS (0-10)** | -3.36 (2.3) | -2.26 (2.7) | -1.1 (-3.5 to 1.3) |
| **Patient global assessment of disease activity VAS (0-10)** | -3.48 (2.5) | -1.36 (2.8) | -2.1 (-4.6 to 0.4) |
| **MMT-8** | 10.14 (19.4) | 12.00 (14.6) | -1.9 (-18.7 to 15.0) |
| **HAQ-DI** | -0.27 (0.5) | -0.09 (0.5) | -0.2 (-0.7 to 0.3) |
| **Muscle Enzyme: CK<sup>1</sup>** | -95.14 (501.7) | 278.14 (365.5) | -373.3 (-830.8 to 84.2) |
| **Muscle Enzyme: AST or ALT<sup>2</sup>** | 3.17 (11.4) | -4.14 (18.4) | 7.3 (-8.3 to 22.9) |
| **Muscle Enzyme: LDH<sup>1</sup>** | -1.4 (109.1) | 55.57 (87.1) | -57.0 (-156.3 to 42.3) |
| **MDAAT** | -0.68 (0.6) | -0.49 (1.1) | -0.2 (-1.1 to 0.7) |
| **CDASI Total Activity Score<sup>3</sup>** | -10.00 (3.7) | -6.00 (8.8) | -4.0 (-11.0 to 3.0) |
| **CDASI Total Damage Score<sup>2</sup>** | 0.33 (4.5) | -0.86 (3.9) | 1.2 (-3.4 to 5.7) |
| **PROMIS PF-20 T-score** | 3.41 (6.0) | -3.08 (7.0) | 6.5 (0.1 to 12.8) |
| **FI-3 score** | 6.43 (12.7) | 1.38 (8.5) | 5.1 (-6.0 to 16.2) |
| **MDI Tool** |  |  |  |
| ***MDI extent of damage score*** | -0.04 (0.1) | -0.00 (0.1) | -0.0 (-0.1 to 0.0) |
| ***MDI severity of damage score*** | -0.10 (0.4) | -0.07 (0.5) | -0.0 (-0.5 to 0.4) |
| ***MDI extended damage score<sup>4</sup>*** | -0.05 (0.1) | -0.03 (0.0) | -0.0 (-0.1 to 0.0) |
| **Perceived Pain** |  |  |  |
| ***Bodily Pain (SF-36)*** | 16.57 (11.3) | -1.63 (31.9) | 18.2 (-5.1 to 41.5) |
| ***HAQ VAS<sup>1</sup>*** | -25.86 (29.8) | -6.16 (25.7) | -19.7 (-47.0 to 7.6) |
| **Fatigue** |  |  |  |
| ***FACIT-Fatigue*** | 6.43 (3.7) | -2.50 (11.6) | 8.9 (0.2 to 17.7) |
| ***Fatigue (SF-36)*** | 13.4 (20.5) | 2.34 (24.3) | 11.0 (-12.0 to 34.1) |
| **Health Status** |  |  |  |
| ***SF-36 MCS<sup>5</sup>*** | 2.56 (9.5) | 2.41 (7.4) | 0.2 (-8.5 to 8.8) |
| ***SF-36 PCS<sup>3</sup>*** | 7.45 (6.2) | -2.04 (13.2) | 9.5 (-0.9 to 19.9) |
| ***EQ-5D-5L Health Utility*** | 0.18 (0.2) | -0.10 (0.3) | 0.27 (0.0 to 0.5) |

Values are presented as mean (SD). <sup>1</sup>One patient did not have the measure assessed at baseline/24 weeks. <sup>2</sup>Two patients have missing data, one in each treatment arm have missing data at baseline and/or 24 weeks. <sup>3</sup>Results based on 12 patients as data missing for 2 patients at baseline and 3 at 24 week visit. <sup>4</sup>Four patients had too much missing data to calculate the MDI extended damage score. <sup>5</sup>One patient did not complete all of the SF-36 questions. Effect estimate was calculated as: mean difference = Immediate start – Delayed-start. Key: VAS = visual analogue scale, MMT = manual muscle testing, HAQ = health assessment questionnaire, CK = creatine kinase, AST = aspartate aminotransferase, ALT = alanine aminotransferase, LDH = lactate dehydrogenase, MDAAT = Myositis Disease Activity Assessment Tool, CDASI = Cutaneous Dermatomyositis Disease Area and Severity Index, PROMIS PF-20 = Patient-Reported Outcomes Measurement Information System - Physical Function 20-item short form, FI-3 = functional index-3, MDI = myositis damage index, SF-36 = Short-Form Health Survey, FACIT = Functional Assessment of Chronic Illness Therapy, MCS = Mental Component Summary, PCS = Physical Component Summary.

**Supplementary Table 3:** Adverse Events at 24 weeks post-active treatment.

| Severity | Treatment arm |  |
| --- | --- | --- |
|  | Immediate-start (N = 20) | Delayed-start (N = 38) |
| G1 – Mild | 11 (55%) | 23 (61%) |
| G2 - Moderate | 6 (30%) | 14 (37%) |
| G3 - Severe | 3 (15%) | 1 (2%) |

## Notes

### Clinical Trial

NCT04208464

### Author Declarations

Research Ethics Committee approval was obtained from UK Health Research Authority (HRA) prior to commencement of the trial (reference 20/NW/0138). Local site permissions were obtained as required in accordance with HRA guidance and institutional requirements. All participants provided informed consent. T

